# Impact Of Covid-19 Pandemic On Mental Health Of Medical Students: A Cross-Sectional Study Using GAD-7 And PHQ-9 Questionnaires

**DOI:** 10.1101/2020.06.24.20138925

**Authors:** Carlos Izaias Sartorão Filho, Wilson Conte de Las Villas Rodrigues, Ricardo Beauchamp de Castro, Arlete Aparecida Marçal, Shirlene Pavelqueires, Luiz Takano, Wilson Luis de Oliveira, Carlos Izaias Sartorão Neto

**Author notes:** **Corresponding author:** Carlos Izaias Sartorão Filho Benedito Spinardi street, 1440 Assis – Sao, Paulo State – Brazil ZIP code: 19815-110.

## Abstract

**Aim:** to evaluate anxiety and depression disorders among medical students during COVID-19 pandemic.

**Methods:** cross-sectional study of medical students conducted in May 2020 with questionnaires regarding social and demographic status and GAD-7 for anxiety and PHQ-9 for depression questionnaires.

**Results:** participated 340 (97.98%) students. Average GAD-7 score was 9.18 (M = 9.18; SD = 4.75); average PHQ-9 score was 12.72 (M = 12.72; SD = 6.62). Results indicate a positive significant relationship between GAD-7 and females, and social distancing affecting finances. Using cut-off score of 10 for GAD-7, 157 (46.17%) students were identified with moderated or severe symptoms of anxiety. For PHQ-9 score, using cut-off of 10, 219 (64.41%) students were identified with moderate or severe symptoms of depression; results indicate a positive significant relationship between PHQ-9 and females and between social distancing affecting finances.

**Conclusion:** analysis demonstrated a higher prevalence of moderated and severe anxiety and depression symptoms among medical students during COVID-19 pandemic, significantly among women and on medical students relating financial impairment related to COVID-19 epidemic.

The project was approved by the Research Ethics Committee of Institution under report number CAAE: 30718220.5.0000.8547

## Introduction and background

The World Health Organization (WHO) declared the novel coronavirus disease 2019 (COVID-19) a pandemic, spreading rapidly and with several risks of morbidity and mortality worldwide. (1) With the intention to prevention, many countries as Brazil introduced restrictions including social distancing, self-isolation, and closure of social and educational institutions. Universities across the world suspended or postponed all activities and substituted the classroom program to online classes.(2) There are a lot of challenges faced by medical schools due to COVID-19 pandemic: shifting from face-to face to online courses, impact on assessments and evaluation programs, travel restrictions among students coming from longer distances, social restrictions, personal financial impairment due to pandemic time, and mental health impact.(2) Emotional disorders in medical undergraduates are common and deserves special attention. (3–7) Psychological reactions to pandemics including maladaptive behaviors, emotional distress and defensive responses deserve special attention in this vulnerable group.(8) The aim of this study is to evaluate the prevalence of anxiety and depression amongst medical students, and the epidemiological, educational and social factors related, during the period of COVID-19 pandemic, in order to identify groups that may require mental health care. The hypothesis that anxiety and depression in medical students are higher during COVID-19 pandemic than related before this period, and is higher among female, in older students, in students not satisfied with on line classes, and in medical students facing financial impairment status during pandemic period.

## Methods

Cross-sectional study including students from the medical school of *Fundação Educacional do Município de Assis* (FEMA), a private school located in the city of Assis, Sao Paulo state, Brazil, applied on May 18 and 19, 2020. The medical course at FEMA is at its 5^th^ year of existence whereas the medical graduation spans six years in Brazil. Inclusion criteria was all students more than 18 years old, officially enrolled in the medical course. Students from FEMA medical school reside nationwide, and at the time of the survey application, they were in social distancing and on stay-at-home recommendation since the postponed presential classes determined by FEMA council on March 17^th^, 2020. The project was approved by the Research Ethics Committee of Institution under report number CAAE: 30718220.5.0000.8547 and all participants were invited to participate after informed consent. The self-reported anonymous on-line survey form was sent via text-message, for each participant, containing informed consent, questionnaires with socio-demographics, educational evaluation and for Anxiety and Depression evaluation to all enrolled medical students of FEMA. Age, gender, and questions about on-line education during pandemic, and question about financial status and its consequences about the continuity on the course were applied. The Generalized Anxiety Disorder (GAD-7) form, designed by Spitzer et al.(9), a seven-item, self-report anxiety questionnaire was applied. The items enquire about the degree to which the patient has been bothered by feeling nervous or anxious, not being able to stop or control worrying, having trouble relaxing, worrying too much about different things, being so restless that it is hard to sit still, becoming easily annoyed and feeling afraid as if something might happen, in the last 2 weeks. We used a version of GAD-7 validated for Portuguese language. (10) We used a cut-off of 10, initially proposed by Kroenke et al (11), with a sensitivity of 89% and specificity of 82% for GAD.

The Patient Health Questionnaire (PHQ) uses scores of the depression criteria classified as “0” (not at all) to “3” (nearly every day). At 9 items, the PHQ depression scale has comparable sensitivity and specificity to other length questionnaires, and consists of the actual 9 criteria upon which the diagnosis of DSM-IV depressive disorders is based. (12) A validated Portuguese version of PHQ-9 was applied. (13) We attempted to determine cut-off of 10 for screening depressive disorder, in accordance with the meta-analysis results from Manea et al (14), that reported no significant differences in pooled sensitivity and specificity for cut-off scores between 8 and 11. A cut-off of 10 is in consonance with the initial validation study, from Kroenke et al, that had a sensitivity of 88% and a specificity of 88% for detecting major depressive disorders. (15)

Collected data were analyzed with SPSS v 20.0 software (IBM Corp., Armonk, NY). Continuous data were expressed in terms of mean (M) and standard deviation (SD). An independent sample t-test was conducted to determine whether there is a difference in GAD-7 score for anxiety and PHQ-9 score for depression between males and females. A one-way ANOVA was conducted to determine the effect of age on scores. A Pearson correlation coefficient was computed to determine the relationship between GAD-7 score, PHQ-9 score, age, gender, and financial status during social isolation period. A value of P<0.05 and confidence interval of 95% were adopted for all analyses.

## Results

Of the total of 347 medical students enrolled, 340 (97.98%) answered the survey. There were 89 male medical students (26.20%) and 251 female medical students (73.80%). One hundred and ten medical students were 18-20 years old (32.40%), 216 were 21-29 years old (63.50%), and 14 were 30 years old or older (4.10%). Declared single status, 317 (93.20%). Five students reported prior suspecting or confirmation of COVID-19 infection (1.47%). 281 (83.14%) students agreed with the stay-at-home order from public health authorities, and 196 (57.82%) declared in total or nearly total isolation at home. Declared afraid of become infected by COVID-19, 288 (84.71%) students. Concerning about on-line classes during pandemic and social distancing, 194 participants (57.06%) declared totally satisfied. When questioned about the income knowledge due to online classes, 294 medical students (86.73%) referred that was less than face-to-face classes. When asked about continuation of postponed educational activities at the campus, 147 (43.24%) responded that prefer continue the online education, 120 (35.29%) responded that prefer suspend the course and 73 (21.47%) were in doubt about continue or not continue the course in this situation. A total of 240 medical undergraduate students responded stay-at-home order affects their financial status and may compromise their continuity on course (70.80%) and 99 students responded that social isolation does not affect their financial status and therefore, their continuity on course (29.20%). (table 1)

**Table 1.** Frequencies for Gender, Age, Marital status, COVID-19 issues, online classes, and financial impairment affecting

| Variable | N | % |
| --- | --- | --- |
| Age |  |  |
| 18-20 | 110 | 32.40 |
| 21-29 | 216 | 63.50 |
| 30 and older | 14 | 4.10 |
| Gender |  |  |
| Male | 89 | 26.20 |
| Female | 251 | 73.80 |
| Marital status |  |  |
| Single | 317 | 93.20 |
| No single | 33 | 6.80 |
| Prior COVID infection or suspected | 05 | 1.47 |
| Social distancing agreement | 196 | 57.82 |
| Afraid of being COVID-19 infection | 288 | 84.71 |
| Satisfaction about online classes | 194 | 57.06 |
| Referred less income knowledge with online classes | 294 | 86.73 |
| Online classes should continue whether stay-at-home status persists | 147 | 43.24 |
| Social distancing and financial impairment may affect their graduation | 240 | 70.80 |

The average GAD-7 score for anxiety was 9.18 (M = 9.18; SD = 4.75) and the average PHQ-9 score for depression was 12.72 (M = 12.72; SD = 6.62). Using a cut-off score of 10 for GAD-7, we found 157 (46.17%) medical students with moderated or severe symptoms of GAD. For PHQ-9 score, using a cut-off of 10, 219 (64.41%) medical students were identified as reporting moderate or severe symptoms of depression. (table 2)

**Table 2.** Descriptive statistics for PHQ-9 score, and for GAD-7 score. N= 340 M: mean, SD: standard deviation, CI: confidence interval

| PHQ-9 | 0 - 4 | 5 - 9 | 10 - 14 | 15 - 19 | 20 - 27 |
| --- | --- | --- | --- | --- | --- |
| N | 35 | 86 | 86 | 63 | 70 |
| % | 10.29 | 25.29 | 25.29 | 18.53 | 20.59 |
| SD | 24.74 | 60.81 | 60.81 | 44.54 | 49.49 |
| 95% CI | 2.630 | 6.463 | 6.463 | 4.735 | 5.261 |

| GAD-7 | 0 - 5 | 6 -10 | 11 - 15 | 16 -21 |
| --- | --- | --- | --- | --- |
| N | 53 | 157 | 88 | 42 |
| % | 15.59 | 46.18 | 25.88 | 12.35 |
| SD | 1.57 | 1.51 | 1.49 | 0.84 |
| 95% CI | 0.423 | 0.235 | 0.311 | 0.253 |

|  | M | SD | 95% CI |
| --- | --- | --- | --- |
| PHQ-9 | 12.72 | 6.62 | 0.704 |
| GAD-7 | 9.18 | 4.75 | 0.505 |

An independent samples t-test (table 3) was conducted to determine whether there is a difference in GAD-7 score for anxiety between males and females. The results indicate a significant difference between males (M = 8.15; SD = 4.41) and females (M = 9.55; SD = 4.82); [t (338) = -2.41; p = 0.015]. The 95% confidence interval of the difference between means ranged from [-2.55 to -0.26] and does indicate a significant difference between the sample means. We therefore reject the null hypothesis and conclude that there is a difference in GAD-7 score for anxiety between males and females.

**Table 3.**
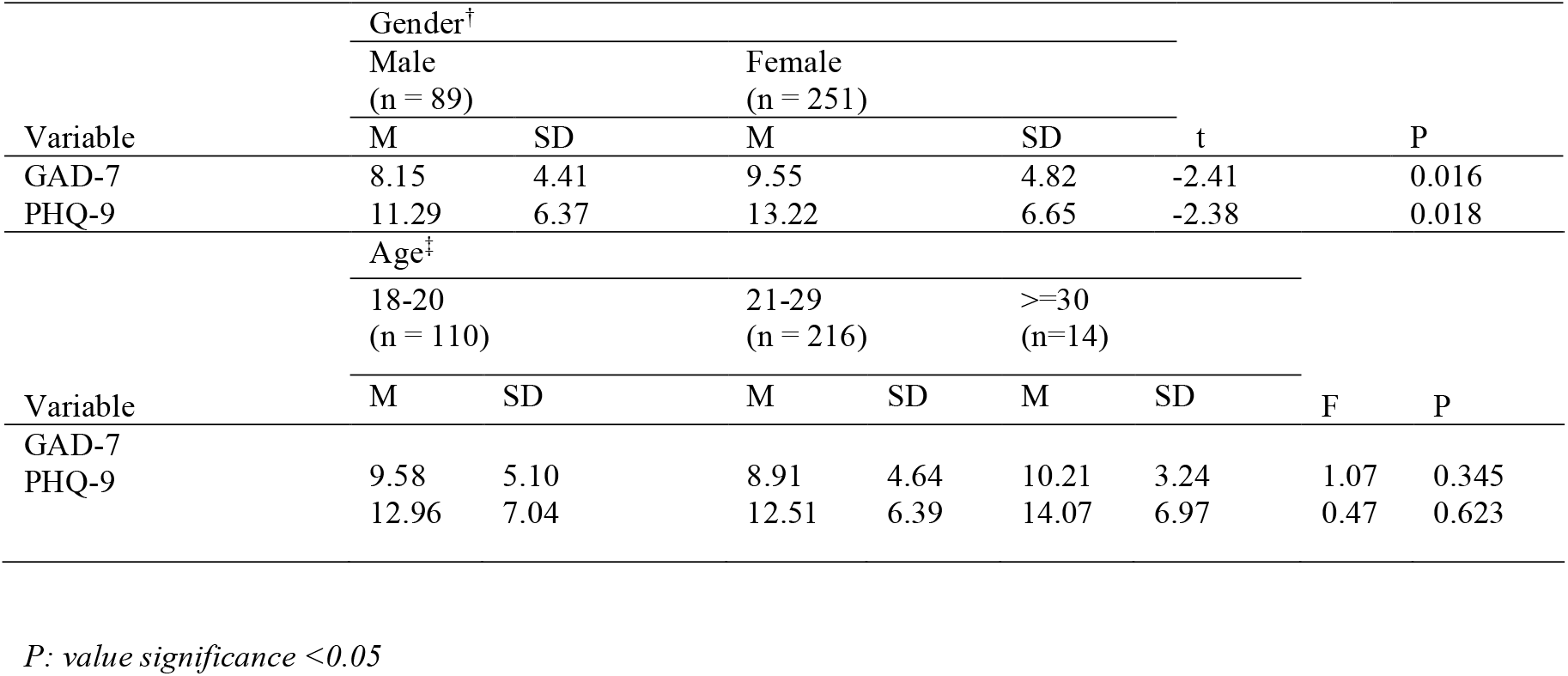
Independent samples t-test results for gender ^†^; one-way ANOVA results for age^‡^. M: mean, SD: standard deviation

| Variable | Gender<sup>†</sup> |  |  |  |  |  |  |  |
| --- | --- | --- | --- | --- | --- | --- | --- | --- |
|  | Male<br>(n = 89) |  | Female<br>(n = 251) |  |  |  |  |  |
|  | M | SD | M | SD | t |  | P |  |
| GAD-7 | 8.15 | 4.41 | 9.55 | 4.82 | -2.41 |  | 0.016 |  |
| PHQ-9 | 11.29 | 6.37 | 13.22 | 6.65 | -2.38 |  | 0.018 |  |
| Variable | Age<sup>‡</sup> |  |  |  |  |  |  |  |
|  | 18-20<br>(n = 110) |  | 21-29<br>(n = 216) |  | >=30<br>(n=14) |  |  |  |
|  | M | SD | M | SD | M | SD | F | P |
| GAD-7 | 9.58 | 5.10 | 8.91 | 4.64 | 10.21 | 3.24 | 1.07 | 0.345 |
| PHQ-9 | 12.96 | 7.04 | 12.51 | 6.39 | 14.07 | 6.97 | 0.47 | 0.623 |

*P*: value significance <0.05

An independent samples t-test was conducted to determine whether there is a difference in PHQ-9 score for depression between males and females. The results indicate a significant difference between males (M = 11.29; SD = 6.37) and females (M = 13.22; SD = 6.65); [t (338) = -2.38; p = 0.018]. The 95% confidence interval of the difference between means ranged from [-3.52 to -0.33] and does indicate a significant difference between the sample means. We therefore reject the null hypothesis and conclude that there is a difference in PHQ-9 score for depression between males and females.

A one-way ANOVA (table 3) was conducted to determine the effect of age (18-20; 21-26; >=30) on GAD-7 score for anxiety. The results indicate a non-significant effect, F (2, 337) = 1.07, p = 0.345. We therefore fail to reject the null hypothesis that the different levels of age have the same effect on GAD-7 score for anxiety. A one-way ANOVA was conducted to determine the effect of age (18-20; 21-26; >=30) on PHQ-9 score for depression. The results indicate a non-significant effect, F (2, 337) = 0.47, p = 0.623. We therefore fail to reject the null hypothesis that the different levels of age have the same effect on PHQ-9 score for depression.

A Pearson correlation coefficient (table 4) was computed to determine the relationship between GAD-7 total score and age 18-20; GAD-7 total score and age 21-29; GAD-7 total score and 30 years and over; GAD-7 total score and female; GAD-7 total score and social distancing does not affect the financial status; does not affect continuity on course; GAD-7 total score and social distancing will affect my financial status and my continuity on course; GAD-7 total score and social distancing may partially affect my financial status and continuity on the course.

**Table 4.** Correlations between GAD-7 and PHQ-9 total score, age, gender, financial status, and continuity on course. Pearson correlation coefficient. N= 340

|  |  | Total score | Age 18 - 20 | Age 21-29 | Age >=30 | Female | † | ‡ | § |
| --- | --- | --- | --- | --- | --- | --- | --- | --- | --- |
| **GAD-7** | r | 1 | 0.058 | -0.075 | 0.045 | 0.130* | -0.114* | 0.150** | -0.002 |
|  | P |  | 0.284 | 0.167 | 0.407 | .016 | 0.036 | 0.005 | 0.972 |
| Age 18 - 20 | r | 0.058 | 1 | -0.913** | -0.143** | 0.126* | 0.058 | -0.101 | 0.038 |
|  | P | 0.284 |  |  |  | 0.020 | 0.288 | 0.063 | 0.484 |
| Age 21-29 | r | -0.075 | -0.913** | 1 | -.274** | -0.118* | -0.028 | 0.063 | -0.036 |
|  | P | 0.167 |  |  |  | 0.030 | 0.607 | 0.248 | 0.514 |
| Age >=30 | r | 0.045 | -0.143** | -0.274** | 1 | -0.011 | -0.068 | 0.085 | -0.004 |
|  | P | 0.407 |  |  |  | 0.836 | 0.211 | 0.116 | 0.946 |
| Female | r | 0.130* | 0.126* | -0.118* | -0.011 | 1 | 0.044 | -0.029 | 0.004 |
|  | P | 0.016 | 0.020 | 0.030 | 0.836 |  | .418 | 0.592 | 0.944 |
| **PHQ-9** | r | 1 | 0.025 | -0.042 | 0.043 | 0.128* | -0.077 | 0.162** | -0.050 |
|  | P |  | 0.645 | 0.441 | 0.434 | 0.018 | 0.155 | 0.003 | 0.354 |
| Age 18 - 20 | r | 0.025 | 1 | -0.913** | -0.143** | 0.126* | 0.058 | -0.101 | 0.038 |
|  | P | 0.645 |  |  |  | 0.020 | 0.288 | 0.063 | 0.484 |
| Age 21-29 | r | -0.042 | -0.913** | 1 | -0.274** | -0.118* | -0.028 | 0.063 | -0.036 |
|  | P | 0.441 |  |  |  | 0.030 | 0.607 | 0.248 | 0.514 |
| Age >=30 | r | 0.043 | -0.143** | -0.274** | 1 | -0.011 | -0.068 | 0.085 | -0.004 |
|  | P | 0.434 |  |  |  | 0.836 | 0.211 | 0.116 | 0.946 |
| Female | r | 0.128* | 0.126* | -0.118* | -0.011 | 1 | 0.044 | -0.029 | 0.004 |
|  | P | 0.018 | 0.020 | 0.030 | 0.836 |  | 0.418 | 0.592 | 0.944 |

† Social distancing does not affect my financial status. Does not affect continuity on course.
‡ Social distancing affects my financial status. Does affect my continuity on course.
§ Social distancing may partially affect my financial status. Does affect my continuity on the course.
\* Correlation is significant at the 0.05 level (2-tailed).
\*\* Correlation is significant at the 0.01 level (2-tailed).

The results indicate: a positive non-significant relationship between GAD-7 total score and age 18-20; r (340) = 0.058, p = 0.284. A negative non-significant relationship between GAD-7 total score and age 21-29; r (340) = -0.075, p = 0.167. A positive non-significant relationship between GAD-7 total score and age 30 and over; r (340) = 0.045, p = 0.407. A positive significant relationship between GAD-7 total score and female; r (340) = 0.130, p = 0.016 < 0.05. A negative significant relationship between GAD-7 total score and social distancing does not affect my financial status. Does not affect continuity on course; r (339) = -0.114, p = 0.036 < 0.05. A positive significant relationship between GAD-7 total score and social distancing will affect my financial status and my continuity on course; r (340) = 0.150, p = 0.005 < 0.05. A negative non-significant relationship between GAD-7 total score and social distancing may partially affect my financial status and continuity on the course; r (340) = -0.002 p = 0.972.

A Pearson correlation coefficient was computed to determine the relationship between PHQ-9 total score and age 18-20; PHQ-9 total score and age 21-29; PHQ-9 total score and 30 years and over; PHQ-9 total score and female; PHQ-9 total score and social distancing does not affect my financial status. Does not affect continuity on course; PHQ-9 total score and social distancing will affect my financial status and my continuity on course; PHQ-9 total score and social distancing may partially affect my financial status and continuity on the course. (Table 4). The results indicate: a positive non-significant relationship between PHQ-9 total score and age 18-20; r (340) = 0.025, p = 0.645. A negative non-significant relationship between PHQ-9 total score and age 21-29; r (340) = -0.042, p = 0.441. A positive non-significant relationship between PHQ-9 total score and age 30 and over; r (340) = 0.043, p = 0.434. A positive significant relationship between PHQ-9 total score and female; r (340) = 0.128, p = 0.018 < 0.05. A negative non-significant relationship between PHQ-9 total score and social distance does not affect my financial status. Does not affect continuity on course; r (339) = -0.077, p = 0.155. A positive significant relationship between PHQ-9 total score and social distancing will affect my financial status and my continuity on course; r (340) = 0.162, p = 0.003 < 0.05. A negative non-significant relationship between PHQ-9 total score and social distancing may partially affect my financial status and continuity on the course; r (340) = -0.050 p = 0.354.

Tables 5 and 6 show individualized descriptive statistics for GAD-7 and PHQ-9, respectively.

**Table 5.**
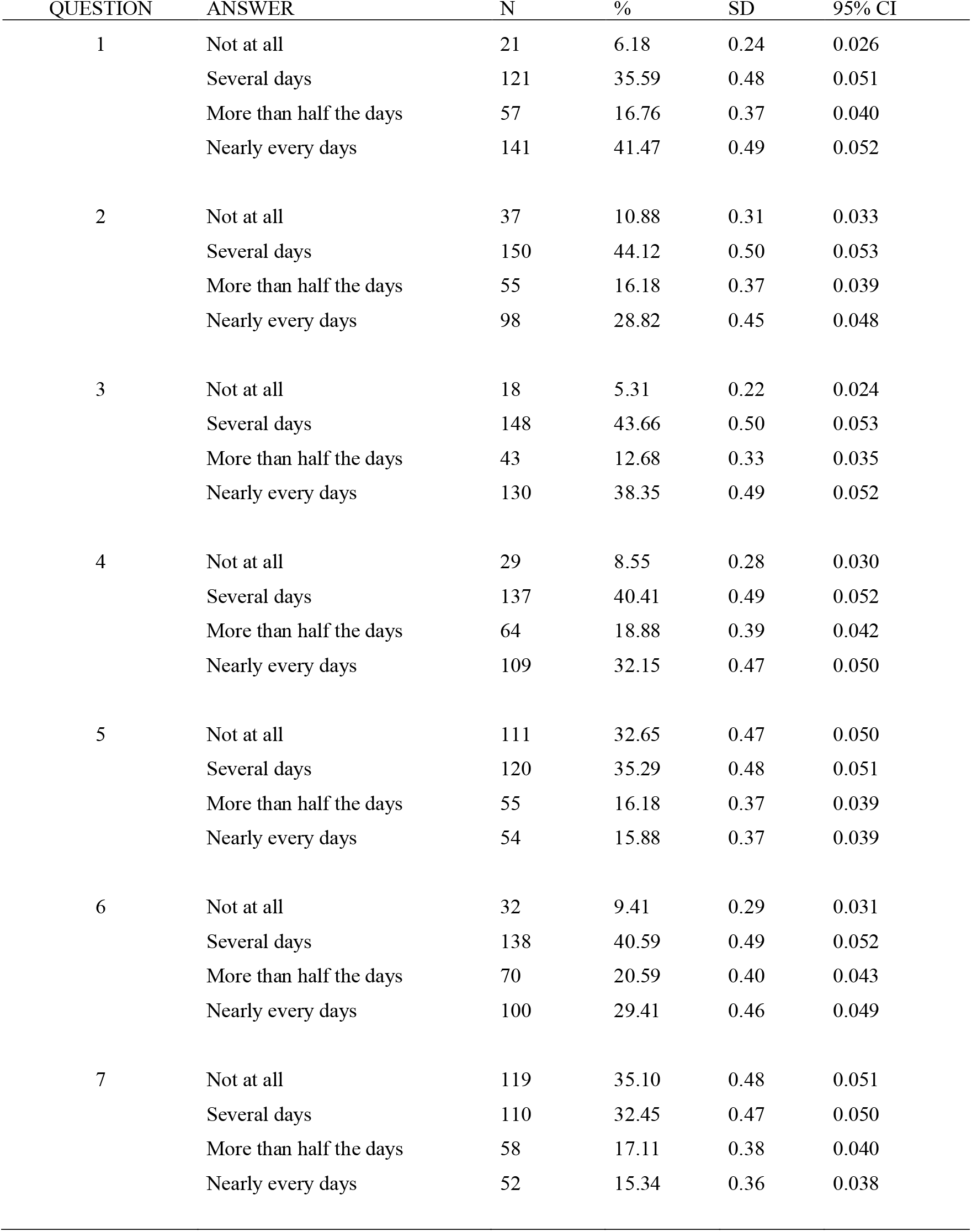
GAD-7 descriptive statistics. N=340 SD: standard deviation, CI: confidence interval

| QUESTION | ANSWER | N | % | SD | 95% CI |
| --- | --- | --- | --- | --- | --- |
| 1 | Not at all | 21 | 6.18 | 0.24 | 0.026 |
|  | Several days | 121 | 35.59 | 0.48 | 0.051 |
|  | More than half the days | 57 | 16.76 | 0.37 | 0.040 |
|  | Nearly every days | 141 | 41.47 | 0.49 | 0.052 |
| 2 | Not at all | 37 | 10.88 | 0.31 | 0.033 |
|  | Several days | 150 | 44.12 | 0.50 | 0.053 |
|  | More than half the days | 55 | 16.18 | 0.37 | 0.039 |
|  | Nearly every days | 98 | 28.82 | 0.45 | 0.048 |
| 3 | Not at all | 18 | 5.31 | 0.22 | 0.024 |
|  | Several days | 148 | 43.66 | 0.50 | 0.053 |
|  | More than half the days | 43 | 12.68 | 0.33 | 0.035 |
|  | Nearly every days | 130 | 38.35 | 0.49 | 0.052 |
| 4 | Not at all | 29 | 8.55 | 0.28 | 0.030 |
|  | Several days | 137 | 40.41 | 0.49 | 0.052 |
|  | More than half the days | 64 | 18.88 | 0.39 | 0.042 |
|  | Nearly every days | 109 | 32.15 | 0.47 | 0.050 |
| 5 | Not at all | 111 | 32.65 | 0.47 | 0.050 |
|  | Several days | 120 | 35.29 | 0.48 | 0.051 |
|  | More than half the days | 55 | 16.18 | 0.37 | 0.039 |
|  | Nearly every days | 54 | 15.88 | 0.37 | 0.039 |
| 6 | Not at all | 32 | 9.41 | 0.29 | 0.031 |
|  | Several days | 138 | 40.59 | 0.49 | 0.052 |
|  | More than half the days | 70 | 20.59 | 0.40 | 0.043 |
|  | Nearly every days | 100 | 29.41 | 0.46 | 0.049 |
| 7 | Not at all | 119 | 35.10 | 0.48 | 0.051 |
|  | Several days | 110 | 32.45 | 0.47 | 0.050 |
|  | More than half the days | 58 | 17.11 | 0.38 | 0.040 |
|  | Nearly every days | 52 | 15.34 | 0.36 | 0.038 |

**Table 6.** PHQ-9 descriptive statistics. N=340 SD: standard deviation, CI: confidence interval

| QUESTION | ANSWER | N | % | SD | 95%CI |
| --- | --- | --- | --- | --- | --- |
| 1 | Not at all | 39 | 11.54 | 0.319 | 0.034 |
|  | Several days | 98 | 28.99 | 0.454 | 0.048 |
|  | More than half the days | 68 | 20.12 | 0.401 | 0.043 |
|  | Nearly every days | 133 | 39.35 | 0.489 | 0.052 |
| 2 | Not at all | 63 | 18.53 | 0.389 | 0.041 |
|  | Several days | 112 | 32.94 | 0.471 | 0.050 |
|  | More than half the days | 72 | 21.18 | 0.409 | 0.043 |
|  | Nearly every days | 93 | 27.35 | 0.446 | 0.047 |
| 3 | Not at all | 55 | 16.18 | 0.369 | 0.039 |
|  | Several days | 74 | 21.76 | 0.413 | 0.044 |
|  | More than half the days | 63 | 18.53 | 0.389 | 0.041 |
|  | Nearly every days | 148 | 43.53 | 0.497 | 0.053 |
| 4 | Not at all | 24 | 7.06 | 0.257 | 0.027 |
|  | Several days | 103 | 30.29 | 0.460 | 0.049 |
|  | More than half the days | 72 | 21.18 | 0.409 | 0.043 |
|  | Nearly every days | 141 | 41.47 | 0.493 | 0.052 |
| 5 | Not at all | 77 | 22.65 | 0.419 | 0.045 |
|  | Several days | 93 | 27.35 | 0.446 | 0.047 |
|  | More than half the days | 63 | 18.53 | 0.389 | 0.041 |
|  | Nearly every days | 107 | 31.47 | 0.465 | 0.049 |
| 6 | Not at all | 111 | 32.65 | 0.470 | 0.050 |
|  | Several days | 91 | 26.76 | 0.443 | 0.047 |
|  | More than half the days | 52 | 15.29 | 0.360 | 0.038 |
|  | Nearly every days | 86 | 25.29 | 0.435 | 0.046 |
| 7 | Not at all | 60 | 17.54 | 0.382 | 0.041 |
|  | Several days | 149 | 43.57 | 0.497 | 0.053 |
|  | More than half the days | 41 | 11.99 | 0.326 | 0.035 |
|  | Nearly every days | 92 | 26.90 | 0.445 | 0.047 |
| 8 | Not at all | 182 | 53.53 | 0.499 | 0.053 |
|  | Several days | 86 | 25.29 | 0.435 | 0.046 |
|  | More than half the days | 43 | 12.65 | 0.333 | 0.035 |
|  | Nearly every days | 29 | 8.53 | 0.280 | 0.030 |
| 9 | Not at all | 294 | 86.47 | 0.343 | 0.036 |
|  | Several days | 23 | 6.76 | 0.252 | 0.027 |
|  | More than half the days | 10 | 2.94 | 0.169 | 0.018 |
|  | Nearly every days | 13 | 3.82 | 0.192 | 0.020 |

## Discussion

We demonstrated that 157 (46.17%) of medical students scored 10 or more, as having symptoms moderated or severe symptoms of anxiety in GAD-7 questionnaire and 219 (64,41%) scored as having moderated or severe symptoms of depression.

Another study conducted in Brazil showed 38.2% of symptoms of depression on medical students (16). Moutinho in 2017 reported 34.6% of depressive symptom and 37.2% of anxiety symptoms in Brazilian medical students.(5) Puthran et al in 2016 (7) published a meta-analysis finding for a total of 62728 medical students across 77 studies, a global prevalence of depression among medical students of 28.0% (95% confidence interval (CI) 24.2-32.1). In that meta-analysis, women were more likely to be depressed, but not significantly. Our results revealed a higher prevalence of anxiety and depression among medical students, with statistically significant higher prevalence in female and in the students with alleged impairment of financial status during social distancing period. Studies demonstrated that women have higher prevalence of anxiety and depression, among general population as well as among medical students.(5,17). In India, Iqbal et al reported higher scores of depression, anxiety and stress associated with female gender, lower semester of course, younger age and non-smokers. (18)

Strengths of this study: we used a type of methodology that has been applied in several other studies on medical schools. Furthermore, GAD-7 and PHQ-9 are well known methods of assessment of symptoms of anxiety and depression, respectively. Other strengths were the massive participation of students (97.98%), responding the text message in only 2 days, revealing the compliance with this evaluation. The relevance of studying mental health during the COVID-19 outbreak has been recognized, especially in a population of medical students, that is known more vulnerable to depression, anxiety, and other stress-related conditions. (19)

### Limitations

first, we cannot explain the heterogeneity between other studies, and caution should be taken when interpreting the results. Although the psychometric properties of the GAD-7 were strong, the measure may better serve as a indicator of GAD severity than a screening tool for the presence or absence of GAD. (20) Future studies should investigate the convergent and discriminant validity of the GAD-7 with respect to other criteria (e.g., behavioral, biological, information-processing) that are relevant to the psychopathology of GAD. (20) Second limitation is PHQ-9 is useful only for screening purposes for “current major depressive episode” as a result of its low positive predictive value. (21)

Third, we had only 14 students with 30 years old or older, and this is a possible limitation concerning age sampling. Furthermore, we do not consider another variable analysis, like marital status, race, employment status, because the majority enrolled were single, Caucasian, not worker and financially dependent, according to FEMA academic registry.

Another limitation is that we do not have a baseline data about mental health disorder regarding medical students of FEMA before COVID-19 pandemic, to be compared. Our quality criteria assessment was not validated to a better understanding about social distancing and financial status of each participant, and in our concerning, this may be a bias, when comparing the results from another cohorts.

Our analyses demonstrated a higher prevalence of moderated and severe anxiety and depression symptoms among medical students during COVID-19 pandemic. The results were significantly higher among females, and in medical students that self-reported that social distancing will affect their financial status and, therefore, their continuity on graduation course.

Financial impairment related due COVID-19 epidemic may be considered as a factor for greater rates of anxiety and depression in medical students from a private school. This must be discussed among authorities to guarantee economic stability and to afford the continuity in the course. In our study, we found a significant impact of financial factor in the prevalence of anxiety and depression symptoms.

Interventions regarding mental health in undergraduate medical students, especially in times of COVID-19 pandemic, where stressful environment causes a negative effect on the academic performance, physical health, psychosocial wellbeing, and financial status, is demanded and imperative. The current findings suggest that medical schools and health authorities should offer prevention, early detection, and interventions for mental health disorders in medical undergraduate students.

## Data Availability

The data that support the findings of this study are available from the corresponding author, [author initials], upon reasonable request.

## Disclosure Statement

the authors declare no conflicts of interest

Ethical approval: CAAE: 30718220.5.0000.8547

